# Multimodal LLMs for Retinal Disease Diagnosis via OCT: Few-Shot vs Single-Shot Learning

**DOI:** 10.1101/2024.11.02.24316624

**Authors:** Reem Agbareia, Mahmud Omar, Ofira Zloto, Benjamin S Glicksberg, Girish N Nadkarni, Eyal Klang

## Abstract

**Background and Aim:** Multimodal large language models (LLMs) have shown potential in processing both text and image data for clinical applications. This study evaluated their diagnostic performance in identifying retinal diseases from optical coherence tomography (OCT) images.

**Methods:** We assessed the diagnostic accuracy of GPT-4o and Claude Sonnet 3.5 using two public OCT datasets (OCTID, OCTDL) containing expert-labeled images of four pathological conditions and normal retinas. Both models were tested using single-shot and few-shot prompts, with an overall of 3088 models’ API calls. Statistical analyses were performed to evaluate differences in overall and condition-specific performance.

**Results:** GPT-4o’s accuracy improved from 56.29% with single-shot prompts to 73.08% with few-shot prompts (p < 0.001). Similarly, Claude Sonnet 3.5 increased from 40.03% to 70.98% using the same approach (p < 0.001). Condition-specific analyses revealed similar trends, with absolute improvements ranging from 2% to 64%. These findings were consistent across the validation dataset.

**Conclusion:** Few-shot prompted multimodal LLMs show promise for clinical integration, particularly in identifying normal retinas, which could help streamline referral processes in primary care. While these models fall short of the diagnostic accuracy reported in established deep learning literature, they offer simple, effective tools for assisting in routine retinal disease diagnosis. Future research should focus on further validation and integrating clinical text data with imaging.

## Introduction

In ophthalmology, AI has shown promise in analyzing imaging data for conditions such as Age-related Macular Degeneration (AMD), Diabetic Retinopathy (DR), and other retinal diseases (1–3). Recently, multimodal Large Language Models (LLMs) have gained attention for their capability to process both textual and visual data, which is essential for interpreting medical images alongside clinical information (4). These systems hold the potential to enhance diagnostic accuracy in a range of image-based tasks within ophthalmology (5).

Retinal diseases, such as AMD, DR, and Central Serous Retinopathy (CSR), are common conditions requiring timely and accurate diagnosis (6,7). Typically, the diagnostic process involves detailed examination by ophthalmologists, using optical coherence tomography (OCT) to capture high-resolution cross-sectional images of the retina (6). OCT is highly efficient in diagnosing and monitoring these conditions, allowing precise visualization of retinal layers and pathologies (6). However, interpreting OCT images demands specialized expertise and can be time-consuming given the increasing patient load in ophthalmic clinics (6). Deep learning models have already proven highly effective in diagnosing retinal diseases using OCT images, achieving expert-level accuracy and high diagnostic performance with Area Under the Receiver Operating Characteristic curve (AUROC) values over 93% in large datasets (8,9).

While deep learning models have shown high diagnostic accuracy, multimodal LLMs present a distinct advantage. They are easier to use, require no complex software setups, support zero-shot and few-shot learning, and seamlessly integrate textual and visual data (10,11). These models could streamline the diagnostic workflow by analyzing OCT images alongside clinical notes, offering diagnostic suggestions that could assist primary care providers in making initial assessments. This approach can potentially reduce the burden on ophthalmologists, allowing for more efficient referrals and enabling quicker access to specialized care for patients with retinal diseases.

“Few-shot” learning is a key technique that has been shown to significantly improve the performance of LLMs (12). In few-shot prompts for vision-based tasks, the model is provided with example images from specific disease categories, allowing it to learn and refine its diagnostic capability based on visual references (12,13). This approach has proven to enhance the accuracy of LLMs, especially in complex multimodal tasks like image interpretation (14).

This study evaluates the diagnostic performance of state-of-the-art multimodal LLMs in classifying retinal diseases from OCT images. We compare “single-shot” and “few-shot” prompts in this task. The focus is on assessing overall and condition-specific accuracy against expert ophthalmologist diagnoses.

## Materials and Methods

### Study Design and Dataset

This study evaluated the diagnostic performance of two multimodal LLMs—OpenAI’s GPT-4o and Anthropic’s Claude Sonnet 3.5—in diagnosing retinal diseases using OCT images (**Figure 1**).

**Figure 1:**
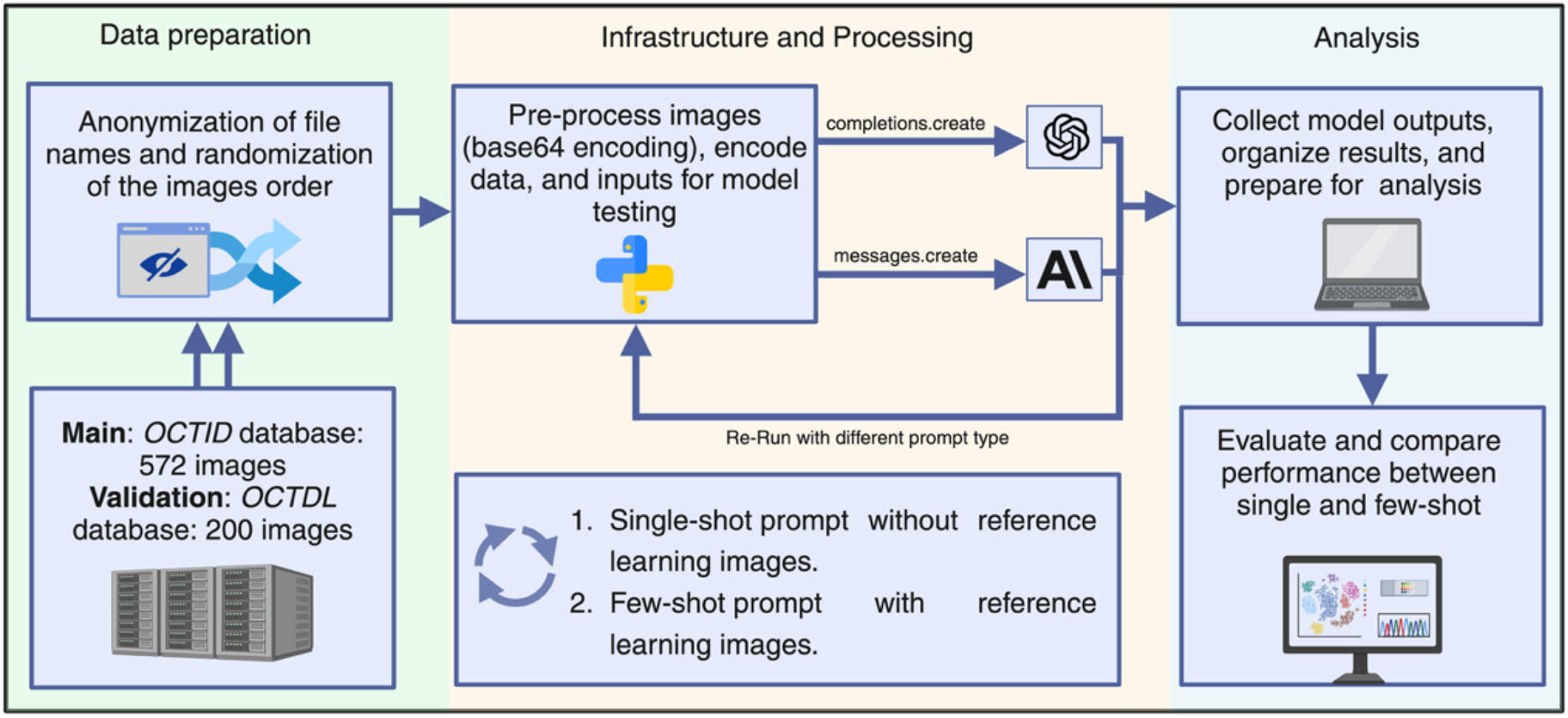
A flowchart summarizing the study’s design.

The study utilized the publicly available OCTID dataset, which contains labeled images of various retinal pathologies, including Age-related Macular Degeneration (AMD), Diabetic Retinopathy (DR), Central Serous Retinopathy (CSR), and Macular Hole (MH). The ground truth labels for these conditions were provided by expert retinal specialists.

The dataset consists of 572 OCT images: 55 AMD, 107 DR, 102 CSR, 102 MH, and 206 normal images, with normal images used for comparison (15).

#### Validation

In addition to the OCTID dataset, we performed a validation step using a random sample of 200 images from the Optical Coherence Tomography Dataset for Image-Based Deep Learning Methods (OCTDL). This validation dataset consists of images labeled according to disease group and retinal pathology. For this study, 67 images of Diabetic Macular Edema (DME), 67 images of AMD, and 66 normal images were randomly selected for validation (16). The selection process ensured random sampling from each category to maintain a balanced representation.

### Model Prompts and Prompt Engineering

To assess the diagnostic accuracy of each model, two types of prompts were utilized:

#### Single-shot Prompt

The models received structured instructions to classify each OCT image into one of the specified retinal conditions. For the OCTID dataset, the models were asked to categorize the images into one of five conditions: AMD, DR, CSR, MH, or Normal. For the validation dataset, the models were asked to categorize the images into one of three conditions: AMD, DME, or Normal.

#### Few-shot Prompt

The models were provided with reference images from each condition category—selected by expert ophthalmologists—alongside the instructions. For the OCTID dataset, reference images for each of the five conditions (AMD, DR, CSR, MH, Normal) were included. For the validation dataset, reference images for three conditions (AMD, DME, Normal) were provided.

Both prompts were designed to mimic real-world diagnostic scenarios in ophthalmology. The full prompt text and the reference images used in the few-shot approach are provided in the **Supplementary Materials Section 1**.

#### Infrastructure

The LLMs were implemented using Python (version 3.9). GPT-4o was accessed via the OpenAI Application Programming Interface (API) using the `completions.create` function, while Claude Sonnet 3.5 was accessed via the Anthropic API using the `messages.create` function. Each OCT image was base64-encoded before being sent to the models through the respective APIs.

To ensure anonymity and randomization, the names of the OCT images’ files were anonymized in the code prior to entry into the models, and the order of images was randomized to prevent any condition-specific ordering or bias. The images were entered consecutively, meaning images of different conditions (AMD, DR, CSR, MH and normal OCT images) were mixed and not analyzed by condition-specific batches.

Each OCT image was classified twice by both models—once using the single-shot prompt and once using the few-shot prompt. A total of 3088 API calls were made across both models.

#### Statistical Analysis

We calculated the means and 95% Confidence Intervals (CI) of the correct diagnoses for each model (GPT-4o and Claude Sonnet 3.5) compared to the ground truth for all conditions. Additionally, we computed the overall performance of each model, combining the results of the single-shot and few-shot iterations. To assess whether the few-shot learning approach led to significant improvements over the single-shot approach, we performed paired t-tests for each model (GPT and Claude), both in general and within each specific condition (AMD, DR, CSR, and MH).

Furthermore, we evaluated the performance of the models on the validation dataset (OCTDL) and compared it to the original dataset (OCTID). Statistical analyses were conducted to check for significant differences in diagnostic accuracy between the two datasets, for each model and across both single-shot and few-shot iterations. This allowed us to assess the generalizability of the model’s performance when applied to different datasets. Statistical significance was set at p < 0.05, and all tests were two-sided. We used R software (version 4.1.2) for the statistical analysis.

## Results

### Overall performance between single-shot and few-shot prompts

The overall performance of the models was evaluated by comparing the mean correct diagnoses across all cases. For GPT, the mean accuracy for single-shot prompts was 56.29% (CI: 52.22% to 60.37%), while for few-shot prompts, the mean accuracy was 73.08% (CI: 69.43% to 76.72%). For Claude, the single-shot performance was lower, with a mean accuracy of 40.03% (CI: 36.01% to 44.06%), and the few-shot performance reached a mean of 70.98% (CI: 67.25% to 74.71%). Both models showed a statistically significant improvement in performance from single-shot to few-shot prompts (p < 0.001 for both GPT and Claude, **Figure 2**).

**Figure 2:**
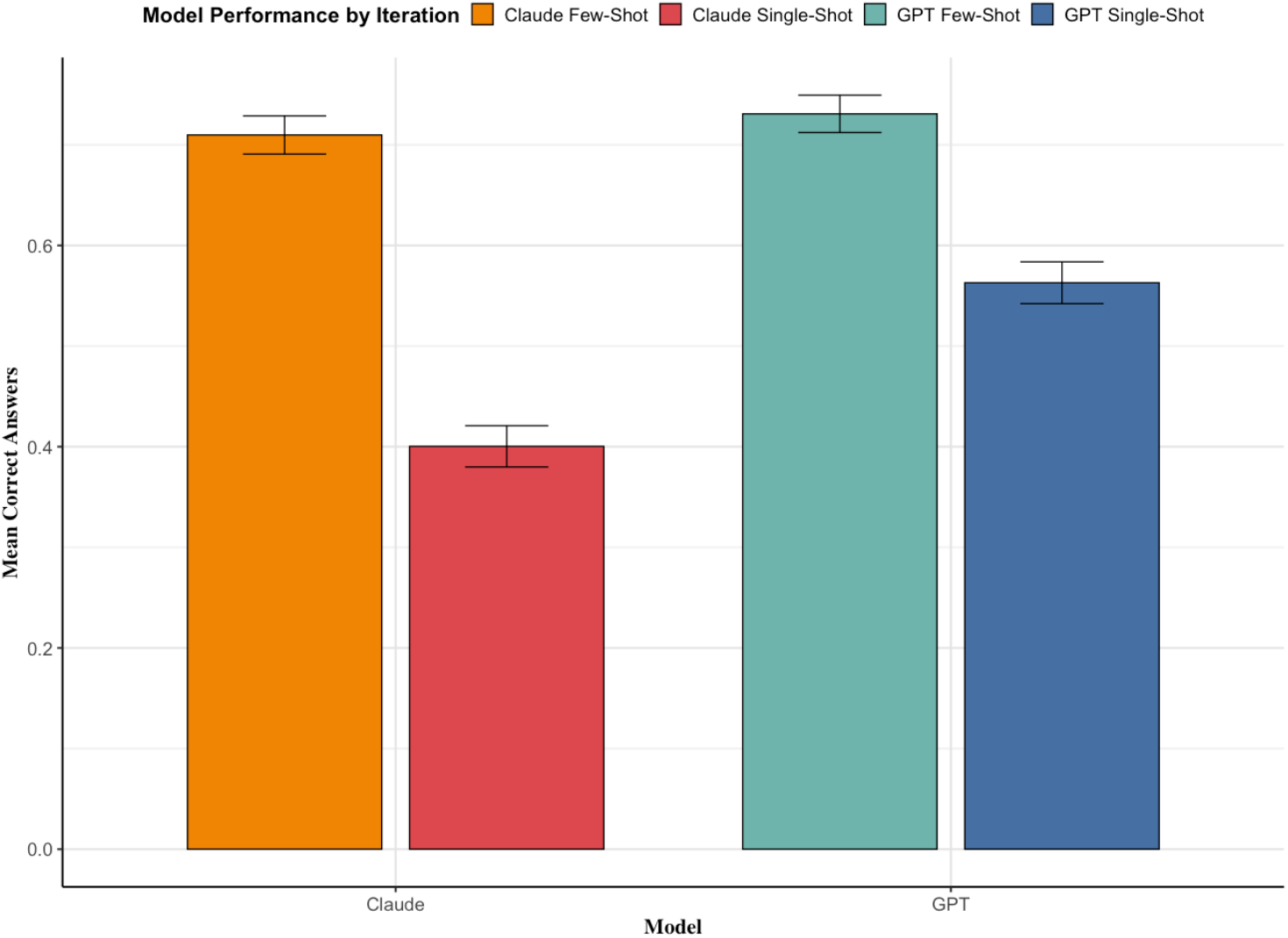
Diagnostic accuracy between single and few shot prompts for the two models overall.

#### Condition-specific performance comparison between few and single-shot prompts

For individual conditions, both GPT and Claude demonstrated performance improvements between single-shot and few-shot learning scenarios, as shown in **Table 1**. For AMD, GPT’s accuracy improved by 29.09% (p < 0.001), and Claude’s accuracy improved by 34.55% (p < 0.001). For CSR, GPT showed a performance gain of 29.41% (p < 0.001), while Claude’s accuracy decreased by 15.69% (p = 0.005). In DR cases, GPT’s accuracy increased by 34.58% (p < 0.001), and although Claude had no correct diagnoses in the single-shot setting, its performance in the few-shot scenario was significant (p < 0.001). For MH, GPT improved by 8.83% (p = 0.006), while Claude improved by 18.63% (p < 0.001). In the normal cases, GPT showed no significant change (p = 0.547), while Claude’s improvement was highly significant, with a 64.08% increase in correct diagnoses (p < 0.001) (**Figure 3**).

**Table 1:** Models performance and improvement across prompts and conditions.

| Condition | Model | Single-shot Performance<br>(CI) | Few-shot Performance (CI) | Improvement<br>(%) | p-value |
| --- | --- | --- | --- | --- | --- |
| AMD | GPT | 18.18% (7.66%, 28.70%) | 47.27% (33.65%, 60.89%) | 29.09% | <0.001 |
|  | Claude | 29.09% (16.70%, 41.48%) | 63.64% (50.51%, 76.76%) | 34.55% | <0.001 |
| CSR | GPT | 45.10% (35.28%, 54.92%) | 74.51% (65.91%, 83.11%) | 29.41% | <0.001 |
|  | Claude | 65.69% (56.32%, 75.06%) | 50.00% (40.13%, 59.87%) | -15.69% | 0.005 |
| DR | GPT | 15.89% (8.85%, 22.93%) | 50.47% (40.84%, 60.10%) | 34.58% | <0.001 |
|  | Claude | 0.00% | 21.50% (13.58%, 29.41%) | 21.50% | <0.001 |
| MH | GPT | 83.33% (75.98%, 90.69%) | 92.16% (86.85%, 97.46%) | 8.83% | 0.006 |
|  | Claude | 70.59% (61.59%, 79.58%) | 89.22% (83.09%, 95.34%) | 18.63% | <0.001 |
| Normal | GPT | 79.61% (74.06%, 85.16%) | 81.55% (76.21%, 86.89%) | 1.94% | 0.547 |
|  | Claude | 35.92% (29.32%, 42.53%) | 100.00% | 64.08% | <0.001 |

**Figure 3:**
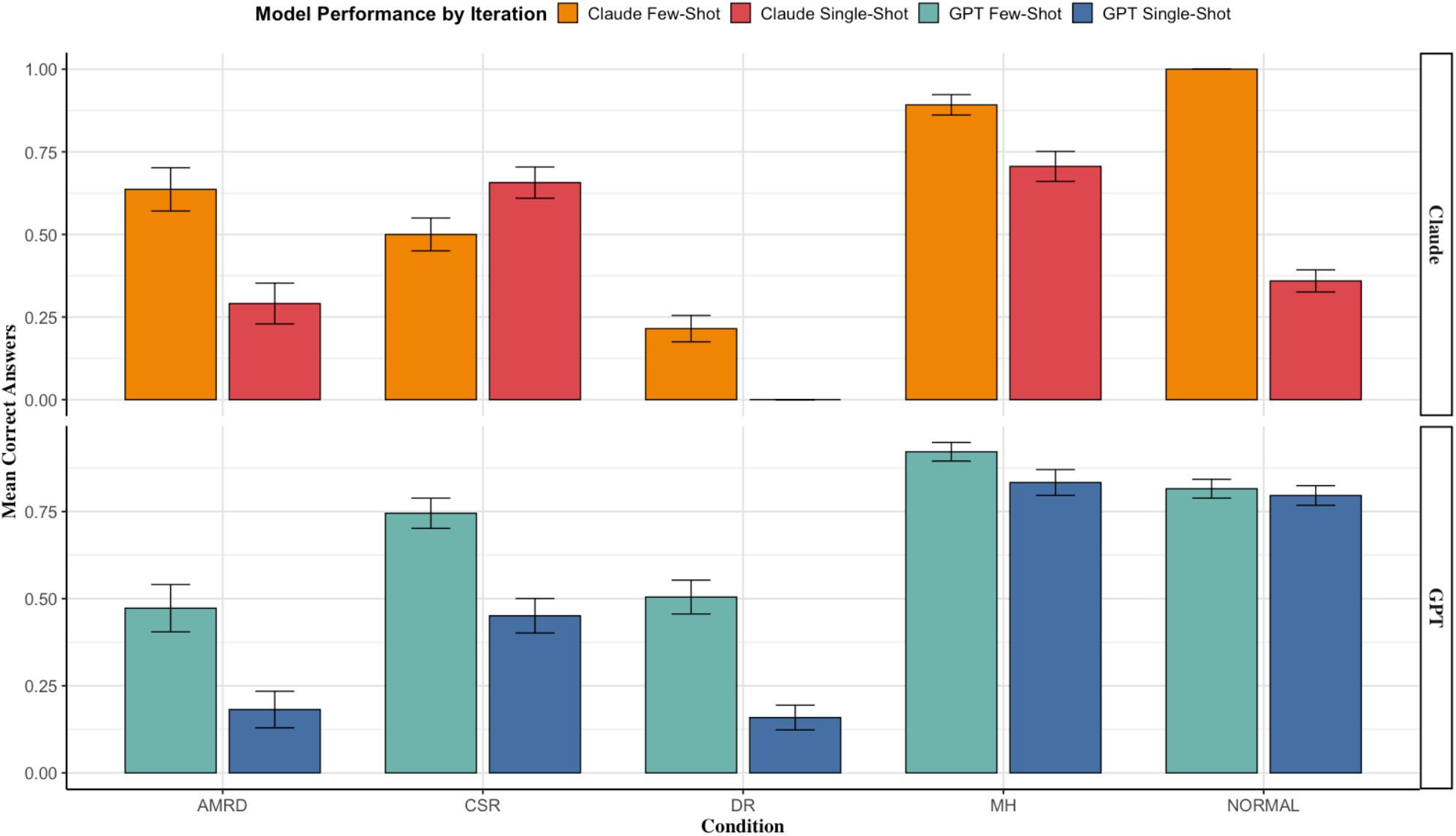
A visual representation of the models’ performances across the prompts and conditions.

#### GPT-4o vs Sonnet 3.5 overall performance across single-shot and few-shot prompts performance

The overall performance of GPT across both few-shot and single-shot conditions was 64.69% (CI: 61.45% to 67.92%), while Claude’s overall performance was 55.51% (CI: 52.39% to 58.63%). A paired t-test comparing the overall performance of GPT and Claude showed a significant difference, with GPT outperforming Claude (p < 0.001).

#### Validation dataset

In the validation dataset, the overall and condition-specific performances for both models improved significantly with few-shot prompts compared to single-shot prompts (**Tables S1 and S2 in the Supplementary Materials, Section 2**). The improvements ranged from 23.4% to 51.5% for GPT-4o, and from 22.5% to 56.3% for Claude Sonnet 3.5, with all differences statistically significant except for Claude in DME (p = 0.118) (**Figure 4**).

**Figure 4:**
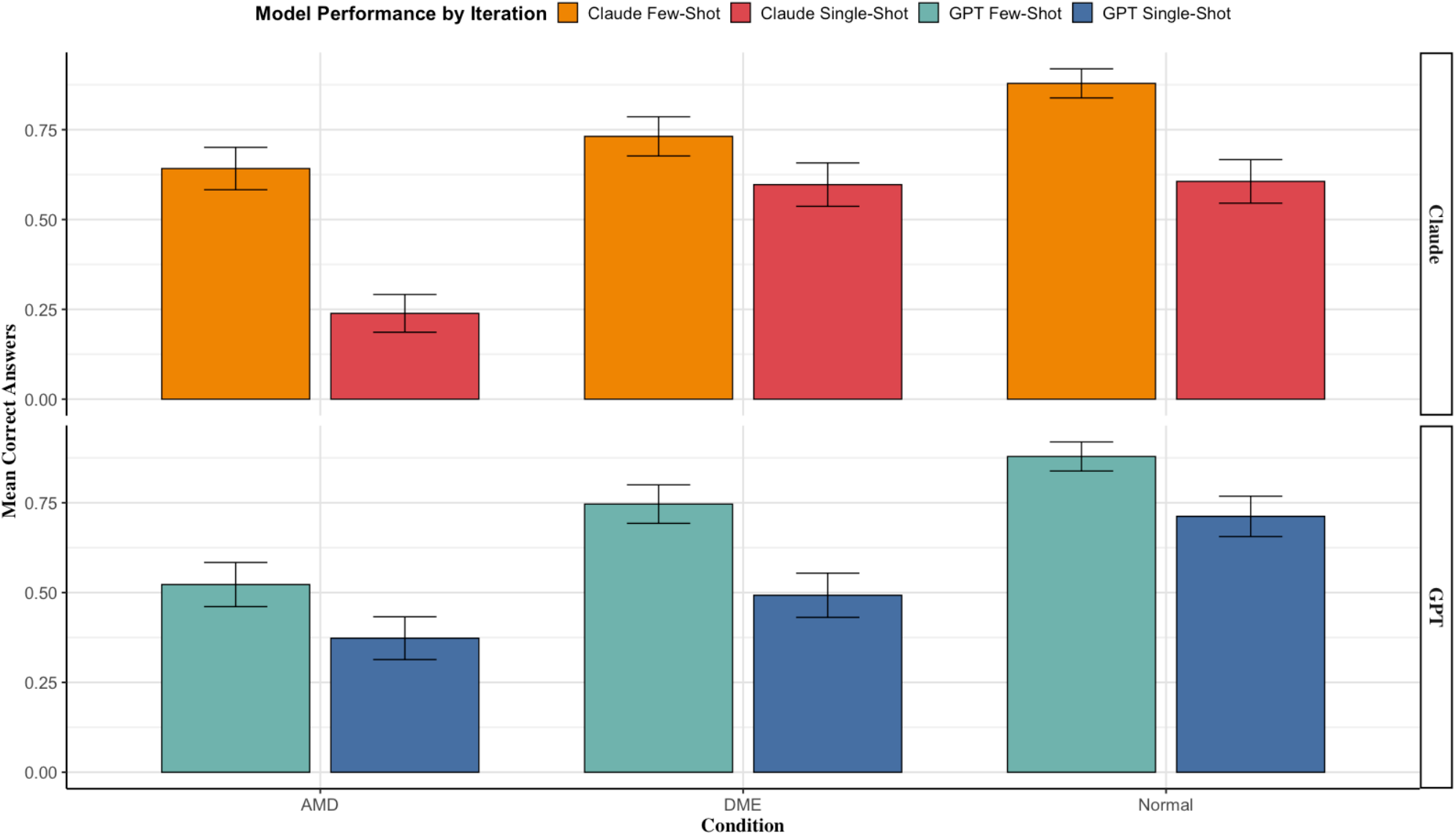
Performance in the validation dataset.

When comparing the test and validation datasets, GPT-4o showed a 2.7% decrease in overall accuracy in the validation dataset (64.7% in the test set vs. 62.0% in the validation, p = 0.048). The single-shot performance of GPT-4o dropped by 3.8% (56.3% vs. 52.5%, p = 0.042), while the few-shot performance showed a 1.6% decrease (73.1% vs. 71.5%, p = 0.065). For Claude Sonnet 3.5, the overall performance improved by 6% in the validation dataset (55.5% in the test set vs. 61.5%, p = 0.032). The single-shot accuracy for Claude increased by 8% (40.0% vs. 48.0%, p = 0.023), and the few-shot accuracy improved by 4% (71.0% vs. 75.0%, p = 0.039) (**Table S3 in the Supplementary Materials, Section 3**).

## Discussion

This study evaluated the diagnostic performance of multimodal LLMs, GPT-4o and Claude Sonnet 3.5, in interpreting retinal OCT images. The results demonstrated a significant improvement in diagnostic accuracy when using few-shot prompts compared to single-shot prompts, with both models showing consistent performance gains across most retinal conditions in two datasets. The effect of using reference images, was crucial in enhancing model accuracy, highlighting its value in multimodal medical image analysis tasks.

GPT-4o’s overall accuracy improved by an average of 16.79%, reaching an accuracy of 73%, with the highest gains seen in DR (34.58%). Claude Sonnet 3.5 demonstrated similar benefits, achieving a notable 64.08% increase in accuracy for identifying normal retinas, bringing its overall accuracy to 70.98%. While Claude’s performance dipped slightly in CSR, the improvement across other conditions remained robust. These findings highlight how few-shot prompts, leveraging visual context, significantly enhance model accuracy, particularly in complex cases such as AMD and DR. This consistent trend not only highlights the capabilities of LLMs in diagnosing OCT images with a reasonable degree of accuracy but also underscores the significant impact of prompt engineering in enhancing the diagnostic performance of multimodal LLMs. These findings align with our previous work, where few-shot prompts with reference images improved LLM diagnostic accuracy in glaucoma detection via fundus images. There, GPT-4o’s accuracy increased by 39.8% and Claude Sonnet 3.5’s by 64.2% (14), further demonstrating how prompt engineering consistently enhances multimodal LLM performance.

Although the accuracy of multimodal LLMs is promising, it falls short compared to the high accuracy typically reported for deep learning (DL)-based models. In our study, few-shot GPT-4o achieved an accuracy of 73.08%, and Claude Sonnet 3.5 reached 70.98%, which is below the 91% to 99% accuracy range often reported in DL models for retinal diseases (17–20). Studies like those by Leandro et al. and Rajagopalan et al. reported accuracies exceeding 97% for conditions such as DME and AMD, highlighting the robustness of DL models, which are optimized for image classification using large datasets (19,20). However, one aspect where our few-shot LLMs show competitive results is in identifying normal retinas. Claude Sonnet 3.5, particularly in the few-shot setting, demonstrated high accuracy in recognizing healthy retinas, comparable to the 93% to 99% range seen in DL studies like Leandro et al.’s work (20). This suggests that few-shot LLMs could be effectively utilized for initial screening of normal retinal cases, potentially streamlining the diagnostic workflow by efficiently ruling out healthy cases in primary care settings.

Despite this, LLMs offer distinct advantages, particularly their multimodal capabilities and ease of use. Unlike deep learning models, which require sophisticated software and large datasets (21), LLMs like GPT-4o and Claude Sonnet 3.5 can be easily deployed in clinical environments without extensive setup (11,22). Their ability to process both clinical notes and visual data, such as intraocular pressure, fundus images, OCT scans, and visual field results, could streamline workflows in primary care. This support can help general practitioners with diagnostic decisions and enable faster referrals to specialists, particularly valuable in resource-limited settings with limited access to ophthalmologists.

Based on our results, multimodal LLMs could assist in clinical settings, particularly in identifying normal optic nerve with high accuracy, which could streamline referrals and reduce the workload on ophthalmologists (23). This would allow specialists to focus on more complex cases. However, while the improvements from prompt engineering are promising, they also highlight the importance of effective interaction with these models. Future healthcare providers may need training on how to craft prompts to maximize the diagnostic potential of LLMs and integrate them into clinical workflows efficiently (24).

This study has limitations. Although we used two datasets with anonymized file names for both testing and validation, we were unable to perform direct comparisons across all specific conditions, as the validation dataset did not include the same five retinal conditions (15,16). Additionally, some of the images in the datasets may have been part of the models’ original training, which could have influenced performance. Finally, our analysis focused solely on imaging, whereas clinical practice typically involves integrating imaging with clinical data (7). Future work should focus on better integration of clinical and imaging data for more comprehensive testing and performance refinement.

In conclusion, the promising results of few-shot prompted multimodal LLMs suggest they could be integrated into clinical practice to streamline and ease the diagnostic process, particularly in primary care settings. While these models still fall short in diagnostic accuracy compared to deep learning techniques, their simplicity and ease of use offer practical solutions for assisting in routine retinal disease diagnoses. Future research should focus on further testing and validation of these models, including fine-tuning, while also exploring the integration of clinical text data with imaging to enhance their diagnostic capabilities and potential clinical utility.

## Data Availability

All data produced in the present study are available upon reasonable request to the authors

## Notes

### Competing Interest Statement

The authors have declared no competing interest.

### Funding Statement

This study did not receive any funding

