## Supplementary Materials for "Multimodal LLMs for Retinal Disease Diagnosis via OCT: Few-Shot vs Single-Shot Learning"

Mount Sinai
Division of Data-Driven and Digital Medicine (D3M), Icahn School of Medicine at Mount Sinai, New York, USA

### Section 1: Prompts

#### Single-shot prompt in-code from the API call of GPT-4o

```
# Structured prompt without showing the image filename
    prompt = """
    As an expert ophthalmologist, your task is to analyze the
provided OCT image. Based on the image, diagnose the patient for
one of the following conditions:
    - 1: Age-related Macular Degeneration (AMD)
    - 2: Diabetic Retinopathy (DR)
    - 3: Central Serous Retinopathy (CSR)
    - 4: Macular Hole (MH)
    - 5: Normal Retina.

    Return your diagnosis in the following structured JSON
format:
    {
      "image_name": "[Anonymous]",
      "diagnosis": [Diagnosis Number]
    }
    Please respond with only the structured JSON, without any
explanation.
    """

    # Call the OpenAI model
    response = client.chat.completions.create(
        model="gpt-4o",
        messages=[
            {"role": "system", "content": "You are an expert
ophthalmologist specializing in retinal OCT diagnosis."},
            {"role": "user", "content": [
                {"type": "text", "text": prompt},
                {"type": "image_url", "image_url": {"url":
f"data:image/jpeg;base64,{image_base64}"}}
            ]}
        ],
        max_tokens=50
    )
```

#### Few-shot prompt in-code from the API call of GPT-4o

```
  # Structured prompt with 5 reference images
    prompt = """
    As an expert ophthalmologist, your task is to analyze the
provided OCT image. Based on the image and the following labeled
reference images, diagnose the patient for one of the following
conditions:
    - 1: Age-related Macular Degeneration (AMD)
    - 2: Diabetic Retinopathy (DR)
    - 3: Central Serous Retinopathy (CSR)
    - 4: Macular Hole (MH)
    - 5: Normal Retina.

    Please consider the following reference images when making
your diagnosis:
    """
    # Add reference images to the prompt (one per condition)
    for ref_image in reference_images:
        prompt += f"- {ref_image['condition']} image data:
data:image/jpeg;base64,{ref_image['encoded_image']}\n"
    # Add the image to be diagnosed
    prompt += """
    Now, based on these reference images, classify the provided
OCT image in the following structured JSON format:
    {
      "image_name": "[Anonymous]",
      "diagnosis": [Diagnosis Number]
    }
    Please respond with only the structured JSON, without any
explanation.
    """

    # Call the OpenAI model
    response = client.chat.completions.create(
        model="gpt-4o",
        messages=[
            {"role": "system", "content": "You are an expert
ophthalmologist specializing in retinal OCT diagnosis."},
            {"role": "user", "content": [
                {"type": "text", "text": prompt},
                {"type": "image_url", "image_url": {"url":
f"data:image/jpeg;base64,{image_base64}"}}
            ]}
        ],
        max_tokens=50
    )
```

### Section 2: Additional tables

#### **Table S1**: Condition-Specific Performance in Few-shot vs Single-shot (Validation Dataset).

| Condition | Model | Mean Single-shot Accuracy | Mean Few-shot Accuracy | % Gain in Few-shot | p-value (Few vs. Single) |
|---|---|---|---|---|---|
| **AMD** | GPT-4o | 52.50% | 71.50% | +40.00% | 0.0491 |
| **AMD** | Claude | 48.00% | 75.00% | +56.25% | < 0.0001 |
| **DME** | GPT-4o | 49.25% | 74.50% | +51.52% | 0.0017 |
| **DME** | Claude | 57.50% | 70.00% | +22.50% | 0.1180 (NS) |
| **Normal** | GPT-4o | 67.00% | 84.00% | +23.40% | 0.0105 |
| **Normal** | Claude | 60.00% | 85.00% | +41.67% | < 0.0001 |

#### **Table S2**: Condition-Specific Performance Comparison Between Test and Validation Datasets

| Condition | Model | Test Dataset Mean (Single + Few) | Validation Dataset Mean (Single + Few) | % Change | p-value |
|---|---|---|---|---|---|
| **AMD** | GPT-4o | 64.00% | 62.00% | -3.13% | 0.0537 (NS) |
| **AMD** | Claude | 57.09% | 61.50% | +7.73% | 0.0462 |
| **DME** | GPT-4o | 65.00% | 74.50% | +14.62% | 0.0248 |
| **DME** | Claude | 59.90% | 70.00% | +16.84% | 0.0411 |
| **Normal** | GPT-4o | 65.50% | 75.50% | +15.27% | 0.0283 |
| **Normal** | Claude | 64.00% | 85.00% | +32.81% | < 0.0001 |

#### **Tables S3**: Comparison of Performance Between Test and Validation Datasets

| Metric | Model | Test Dataset Mean (Single + Few) | Validation Dataset Mean (Single + Few) | % Change (Validation vs. Test) | p-value |
|---|---|---|---|---|---|
| **Overall (Single + Few)** | GPT-4o | 64.69% | 62.00% | -4.15% | 0.0481 |
| **Overall (Single + Few)** | Claude | 55.51% | 61.50% | +10.80% | 0.0323 |
| **Single-shot** | GPT-4o | 56.29% | 52.50% | -6.73% | 0.0427 |
| **Single-shot** | Claude | 40.03% | 48.00% | +19.92% | 0.0239 |
| **Few-shot** | GPT-4o | 73.08% | 71.50% | -2.16% | 0.0655 (NS) |
| **Few-shot** | Claude | 70.98% | 75.00% | +5.66% | 0.0397 |
